# Exploring the possible interventions in children and adolescents with Developmental Coordination Disorder. How far have we come? A Scoping Review

**DOI:** 10.1101/2024.12.31.24319812

**Authors:** Panciroli Maria Chiara, Fiore Simone, Baragetti Andrea, Fascia Matteo

## Abstract

**Background:** Developmental Coordination Disorder (DCD) is a common neurodevelopmental disorder. Children with DCD usually need treatment. Indications for intervention are essentially dependent on the influence of the diagnosis on activities of daily life and the proposed treatments are therefore heterogeneous.

**Objectives:** Identifying and describing the current state of knowledge regarding the treatments available in the literature for the treatment of DCD, defining which of these interventions have been most investigated nowadays, identifying other possible types of intervention that have been introduced in the clinical practice and described by sporadic/anecdotal evidence.

**Study design:** Scoping review

**Eligibility criteria:** This review will take in account studies with any type of research design, with no geographical limit, published after 1994. This criterion was defined as corresponding to the date of publication of the DSM-IV, the first document in chronological order to be considered as a reference for the definition of diagnostic criteria in this scoping review.

Inclusion is limited to articles written in either English or Italian language. The population of interest includes children and adolescents (age 0-18 years old) with a diagnose of Developmental Coordination Disorder (DCD) or a probable-DCD (based on DSM-IV or DSM-IV-TR or DSM-V or ICD-10 or ICD-11 criteria) also in possible co-presence of other neurodevelopmental disorders, with the exception of autism spectrum disorder.

## INTRODUCTION

Developmental Coordination Disorder (DCD) is a common neurodevelopmental disorder, resulting in considerable consequences in daily life; prevalence estimates of 5% to 6% of children are most frequently quoted in the literature (1), with male:female ratios varying from 2:1 to 7:1 (2,3). However, DCD is largely underrecognized by the health care and the educational professionals. (4)

These kids struggle with motor skills, which make them difficult to thrive socially and academically.(2) Research indicates that kids with DCD engage in less physical activity overall, particularly when it comes to team sports.(5) In children with DCD, reduced physical activity has been linked to decreased life satisfaction and poorer self-efficacy. (6,7)

The marked influence of DCD on everyday activities and school performance, and, secondarily, on social participation, physical health, and mental health concerns, combined with the high prevalence rate indicate that the social and economic burden is considerable. (8)

Children with DCD, meeting the diagnostic criteria for DCD, usually need treatment. Indications for intervention are essentially dependent on the influence of the diagnosis on activities of daily life. Approaches to DCD intervention have historically been divided into two main categories: process-oriented approaches, which use activity to target the underlying performance problems and task-oriented approaches, which deal with the performance issue directly.

In the International clinical practice recommendations published in 2019 (8), a new taxonomy for interventions based on ICF terminology was introduced and interventions were therefore grouped into three categories:

1. body function and structure-oriented
2. activity-oriented
3. participation-oriented

In addition, the authors specified that psychosocial factors that may accompany a child’s motor difficulties should be considered when planning intervention. Due to a paucity of comparative studies, the available data on the efficacy of the intervention does not permit precise recommendations for timing, duration, and intensity.

A scoping review on this topic could identify gaps in the literature and highlight areas that require further research.

## OBJECTIVES

Identifying and describing the current state of knowledge regarding the treatments available in the literature for the treatment of DCD in children and adolescents.

Secondary objectives:

- Defining which of these interventions proposed by guidelines have been most investigated in the literature nowadays, the age groups most investigated;
- Identifying other possible types of intervention that have been introduced in the clinical practice and described by sporadic/anecdotal evidence

According to the Joanna Briggs Institute (JBI) guidelines(9,10) a “PCC” strategy was used to state the research question (**Population:** Children and adolescents (0-18 years old) with DCD or those labeled probable-DCD;

; **Concept:** Any types of interventions with children and adolescents with DCD.; **Context:** educational and rehabilitative settings).

## METHODS

### Protocol and registration

Registration on Medxriv

### Eligibility criteria

#### Inclusion criteria

Any type of study design, with no geographical limit, published after 1994. This criterion was defined as corresponding to the date of publication of the DSM-IV, the first document in chronological order to be considered as a reference for the definition of diagnostic criteria in this scoping review.

Inclusion is limited to articles written in either English or Italian language. The population of interest includes children and adolescents (age 0-18 years old) with a diagnose of Developmental Coordination Disorder (DCD) or a probable-DCD (based on DSM-IV(11) or DSM-IV-TR(12) or DSM-5(1) or ICD-10(13) or ICD-11(14) criteria) also in possible co-presence of other neurodevelopmental disorders, except for autism spectrum disorder.

#### Exclusion criteria

All the studies that do not match the PCC described above.

### Information sources

The research will be conducted on the following databases: Medline, Embase, Cochrane Library; We will search for grey literature on Scopus and Web of Science.

### Search Strategy

As recommended in all JBI types of reviews and PRISMA-S(15), a three-step search strategy will be utilized. The first step will be an initial search of an appropriate online database relevant to the topic (PubMed). This initial search will be followed by an analysis of the text words contained in the title and abstract of retrieved papers, and of the index terms used to describe the articles. A background search using all identified keywords and index terms will be undertaken on Pubmed. The reference list of identified reports and articles will be searched for additional sources. No search limitations and filters will be applied except for the language (English and Italian) and the publication year.

Some of the key words used for the creation of the Pumed search string were as follows:

‘developmental coordination disorder’, ‘developmental dyspraxia*’, ‘specific developmental disorder of motor function*’, ‘child’, ‘adolescent’, ‘intervention*’, ‘rehabilitation*’, ‘training’ ‘therap*’. The boolean operators “AND, “OR” and “NOT” were used to refine the search strategy.

### Selection of sources of evidence

Selection process is based on title and abstract by two independent reviewers; disagreements on study selection and data extraction will be discussed with another reviewer if needed.

Selection is performed based on inclusion criteria pre-specified; the software used for the management of the results will be Microsoft Excel and Rayyan. For excluded studies, reasons should be stated on why they were excluded.

There will be a narrative description of the process accompanied by a flowchart of review process (from the PRISMA-ScR statement).

#### Pilot testing of source selectors

Random sample of 25 titles/abstracts is selected, the entire team screens these using the eligibility criteria and definitions/elaboration document, then the team meets to discuss discrepancies and make modifications to the eligibility criteria and definitions/elaboration document. Team only starts screening when 75% (or greater) agreement is achieved. When screening started we used a Microsoft Excel file to manage and catalogue the articles resulting from the search

### Data charting process

Team trial the extraction from one, two or three sources to ensure all relevant results are extracted, by at least two members of the review team. A template data extraction instrument for source details, characteristics and results extraction is provided in appendix.

Pilot step: the extraction form will be tested on two or three sources to ensure all relevant results are extracted, by two blinding members of the review team; Inconsistencies will be resolved by a third reviewer,

This form will be reviewed by the research team and pre-tested by all reviewers before implementation to ensure that the form captures the information accurately, modifications will be detailed in the full scoping review.

Key information’s will be described in a charting table with the description of:

- Author(s)
- Year of publication
- Country of publication
- Study design
- Diagnostic criteria
- Comorbidities
- Aim of the study
- Sample characteristics (age, nationality, N)
- Type of intervention, duration and frequency
- Type of control or other intervention, duration
- Diagnose of the control/ other intervention group
- Outcome measures

### Critical appraisal of individual sources of evidence

No critical appraisal will be performed according to JBI guidelines for Scoping Review.

### Synthesis of results

The results will be presented as a map of the data extracted from the included papers in a diagrammatic, tabular form, and in a descriptive format that aligns with the objectives and scope of the review. Descriptive analysis: distribution of sources of evidence by year or period of publication, countries of origin, area of intervention (clinical, policy, educational, etc.), and research methods. The results can also be classified under the main conceptual categories such as:

- Type of intervention, duration and frequency
- Aim of the study
- Sample characteristics

## Data Availability

All data produced in the present study are available upon reasonable request to the authors

## Notes

### Competing Interest Statement

The authors have declared no competing interest.

### Funding Statement

This study did not receive any funding

